# Electronic Health Record Documentation of Psychiatric Assessments in Massachusetts Emergency Department and Outpatient Settings During the COVID-19 Pandemic

**DOI:** 10.1101/2020.03.30.20048207

**Authors:** Victor M. Castro, Roy H. Perlis

## Abstract

**Question:** How did documentation of psychiatric symptoms in outpatient and emergency room settings change with onset of COVID-19 infection in Eastern Massachusetts?

**Findings:** In this cohort study spanning 2 academic medical centers and 3 community hospitals, prevalence of narrative notes referencing depression or anxiety decreased 75-81% in outpatient settings following onset of coronavirus in March 2019, and by 44–45% in emergency departments.

**Meaning:** The observation that documentation of psychiatric symptoms declined sharply with increasing coronavirus infection in Massachusetts, even as prevalence of such symptoms is anticipated to increase, suggests additional efforts may be required to address these symptoms in the context of COVID-19.

## Introduction

The emergence of the worldwide coronavirus-19 (COVID-19) pandemic has been associated with increased burden of psychiatric symptoms among healthcare workers and the general public,^1,2^ which may increase risk for longer-term sequelae^3,4^. At the same time, the need for quarantine and the strain on clinical resources may reduce the ability of health systems to respond to such symptoms. To quantify shifts in psychiatric evaluation associated with COVID-19, we utilized electronic health records (EHR) from 2 large academic medical centers and 3 affiliated community hospitals. We investigated documentation of psychiatric symptoms among narrative clinical notes as disease activity increased in Eastern Massachusetts.

## Methods

The cohort included all individuals seen in outpatient visits or emergency rooms between January 2, 2019 and March 25, 2020. Sociodemographic data including age, sex, race, and ethnicity were drawn from the Partners Research Patient Data Registry^5^, along with narrative clinical notes. Presence of depression, anxiety, suicide, psychosis, and violence was determined by identifying terms drawn from an expert-curated list associated with NIMH Research Domain Criteria (RDoC)^6^. These terms included depressed, depressive, dysphoric, dysthymic, sad, tearful; anxiety, anxious, fearful, frighten, hypervigilant, nervous, panic, phobia, phobic, scared, stress, tense, worried; suicide, suicidal, suicidality; psychotic, psychosis, hallucination, delusion, paranoid, paranoia, hallucinate, hallucinated, delusional; violence, violent.

Presence of a coronavirus test was determined from the enterprise laboratory feed (LOINC:94309–2). The Partners HealthCare Human Research Committee approved the study protocol.

Primary analysis quantified volume of documentation by individual symptoms by week. Logistic regression was used to examine the relationship between presence of coronavirus testing and psychiatric symptoms, adjusted for age, sex, and race/ethnicity, using R 3.6.0. Notes were treated as clustered within-individual, and limited to those occurring at or before time of testing. STROBE reporting guidelines were followed.

## Results

A total of 205,957 emergency department and 2,483,159 outpatient notes were analyzed (Table 1) representing 60,428 and 541,307 patients, respectively (Supplemental Table 1). Figure 1 illustrates frequency of coronavirus testing (panel A) by week, and frequency of psychiatric terms in notes. The initial 2 decrements in terms correspond to 4-day weeks. Notes with mentions of depression decreased from 1,446 to 816 (i.e., by 44%) for ED and 49,132 to 9,315 (i.e., by 81%) for outpatient from a weekly average in January and February, excluding holidays, to the week spanning 3/19/2020–3/25/2020. Similar patterns were observed for other symptoms.

**Table 1.** Sociodemographic summary of patients seen from 1/2/2020–3/25/2020. Counts and percentages are total number of notes by visit setting.

|  | **ED**<br>**(Notes=205,957)** | **Outpatient**<br>**(Notes=2,483,159)** | **Total**<br>**(Notes=2,689,116)** |
| --- | --- | --- | --- |
| **Gender** |  |  |  |
| Female | 111,679 (54.2%) | 1,525,338 (61.4%) | 1,637,017 (60.9%) |
| **Age (years)** |  |  |  |
| Mean (SD) | 47.1 (23.5) | 52.0 (21.4) | 51.7 (21.663) |
| **Race** |  |  |  |
| Asian | 8,119 (3.9%) | 108,327 (4.4%) | 116,446 (4.3%) |
| Black | 3,0914 (15.0%) | 164,712 (6.6%) | 195,626 (7.3%) |
| Other | 22,224 (10.8%) | 133,952 (5.4%) | 156,176 (5.8%) |
| Unknown | 16,128 (7.8%) | 170,988 (6.9%) | 187,116 (7.0%) |
| White | 128,572 (62.4%) | 1,905,180 (76.7%) | 2,033,752 (75.6%) |
| **Ethnicity** |  |  |  |
| Hispanic | 103,44 (5.0%) | 88,524 (3.6%) | 98,868 (3.7%) |
| **Hospital Type** |  |  |  |
| Academic Medical Centers | 124,518 (60.5%) | 1,920,205 (77.3%) | 2044,723 (76.0%) |
| Community Hospitals | 81,439 (39.5%) | 562,954 (22.7%) | 644,393 (24.0%) |
| **COVID-19 Lab Order** |  |  |  |
|  | 5,635 (2.7%) | 34,165 (1.4%) | 39,800 (1.5%) |
| **Anxiety term in note** |  |  |  |
|  | 39,978 (19.4%) | 978,308 (39.4%) | 1,018,286 (37.9%) |
| **Depression term in note** |  |  |  |
|  | 16,810 (8.2%) | 491,612 (19.8%) | 508,422 (18.9%) |
| **Psychosis term in note** |  |  |  |
|  | 10,272 (5.0%) | 121,612 (4.9%) | 131,884 (4.9%) |
| **Suicide term in note** |  |  |  |
|  | 11,007 (5.3%) | 133,022 (5.4%) | 144,029 (5.4%) |
| **Violence term in note** |  |  |  |
|  | 4,722 (2.3%) | 234,381 (9.4%) | 239,103 (8.9%) |

**Figure 1.**
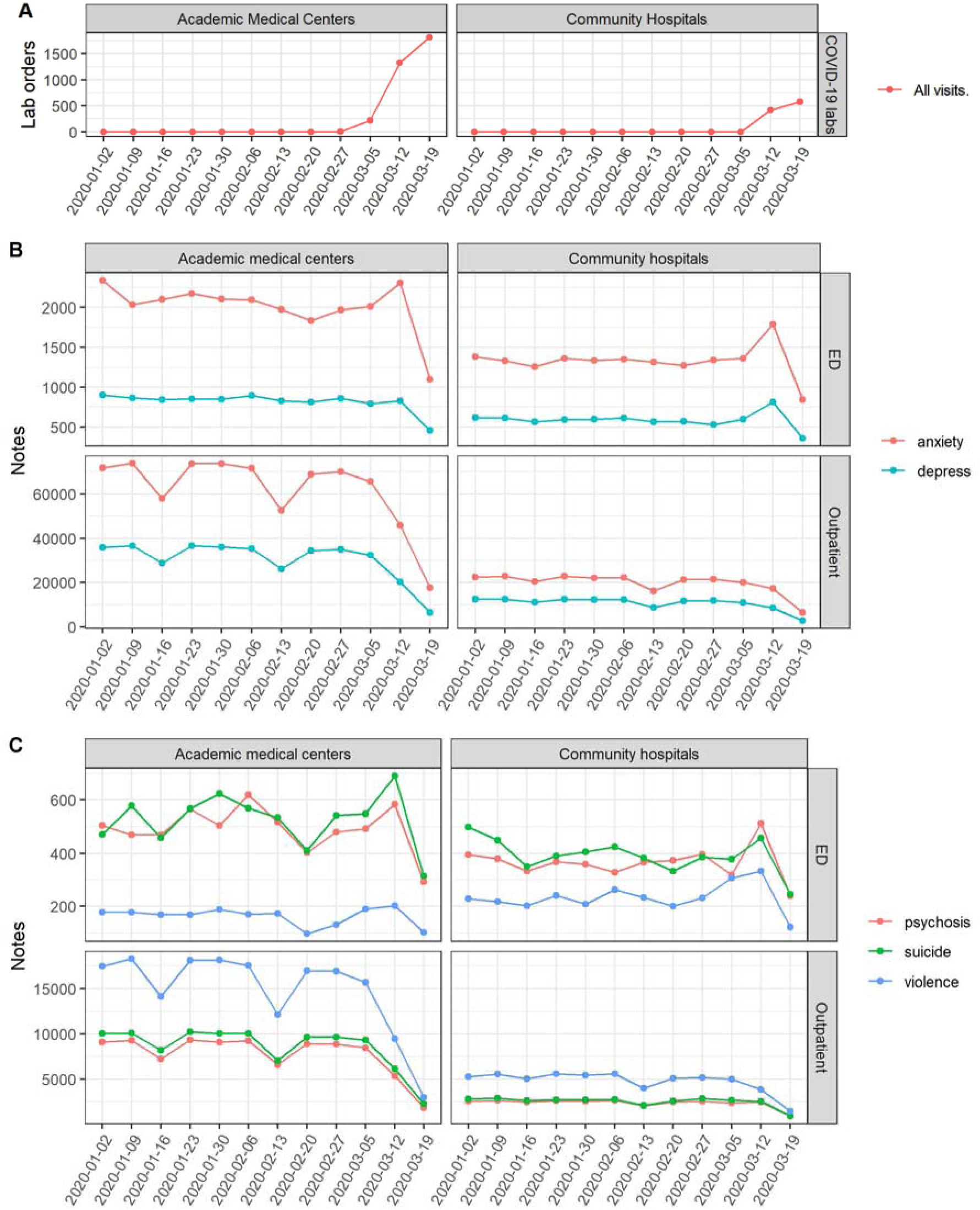
Trends in COVID-19 lab orders and psychiatric tokens by week for date period 1/2/2020 - 3/25/2020. Counts are computed using weeks starting Thursdays and ending on Wednesdays. (A) COVID-19 lab orders from either outpatient or ED patients. (B) Count of notes with anxiety and depression mentions over time. (C) Count of notes with psychosis, suicide, and violence mentions over time.

In the ED setting, where violence was referenced, odds of testing were increased by nearly 50% (OR 1.487, 95% CI 1.249-1.761) (Table 2). In the outpatient setting, notes with presence of psychiatric terms were associated with reduction in likelihood of coronavirus testing for an individual patient (Table 2), with adjusted odds ratios ranging from 0.795 (95% CI 0.770–0.821) for anxiety to 0.631 (95% CI 0.563–0.705) for psychosis.

**Table 2:**
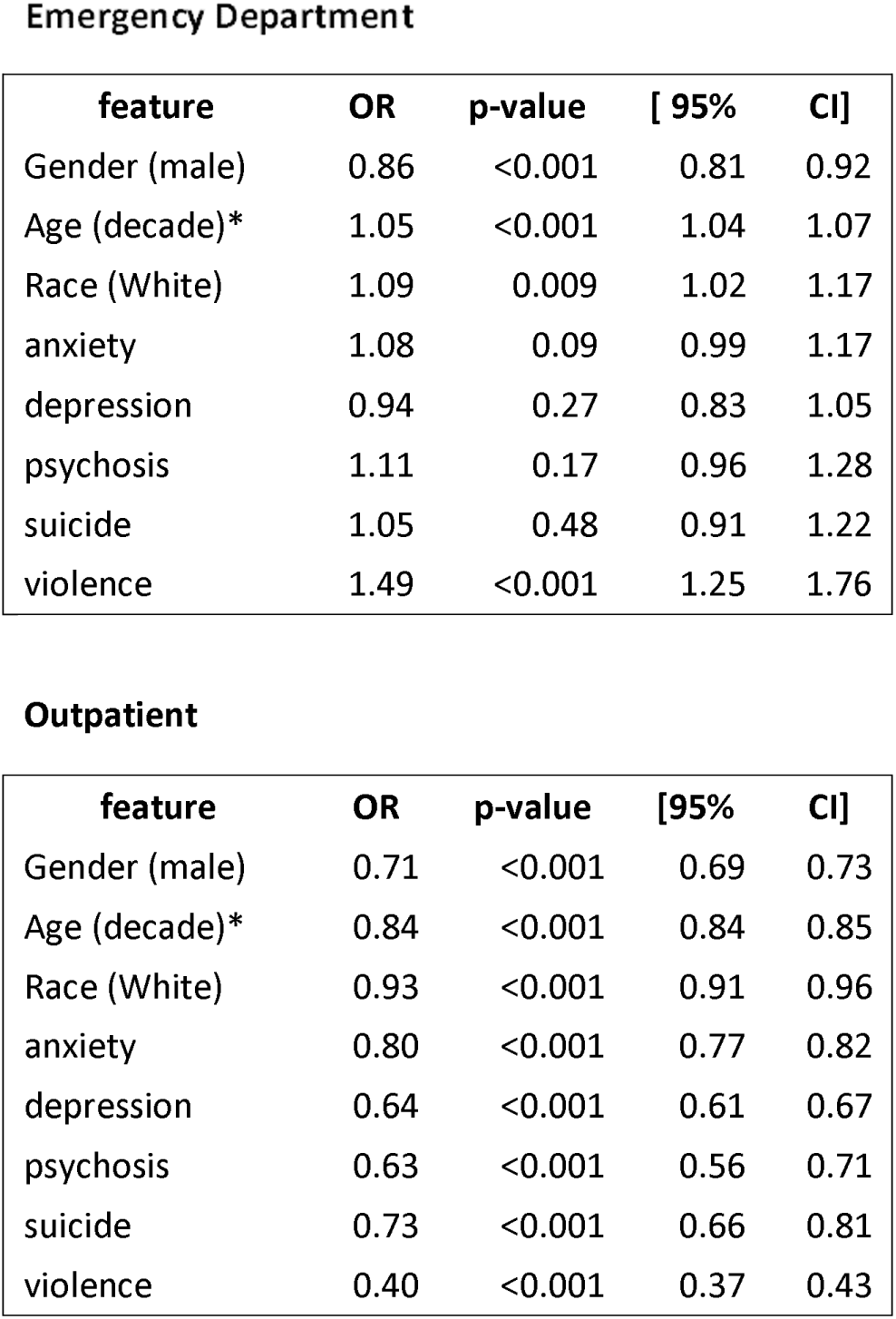
Emergency Department. Clustered logistic regression models examining the relationship between coronavirus lab testing and individual symptoms and individual symptoms. OR, odds ratio; Cl, confidence interval. *For interpretability, age is normalized by subtracting 50 and dividing by 10.

**Emergency Department**
| feature | OR | p-value | [ 95% | CI] |
| --- | --- | --- | --- | --- |
| Gender (male) | 0.86 | <0.001 | 0.81 | 0.92 |
| Age (decade)* | 1.05 | <0.001 | 1.04 | 1.07 |
| Race (White) | 1.09 | 0.009 | 1.02 | 1.17 |
| anxiety | 1.08 | 0.09 | 0.99 | 1.17 |
| depression | 0.94 | 0.27 | 0.83 | 1.05 |
| psychosis | 1.11 | 0.17 | 0.96 | 1.28 |
| suicide | 1.05 | 0.48 | 0.91 | 1.22 |
| violence | 1.49 | <0.001 | 1.25 | 1.76 |

**Table 2:** Clustered logistic regression models examining the relationship between coronavirus lab testing and individual symptoms and individual symptoms. OR, odds ratio; CI, confidence interval. \*For interpretability, age is normalized by subtracting 50 and dividing by 10.
| feature | OR | p-value | [95% | CI] |
| --- | --- | --- | --- | --- |
| Gender (male) | 0.71 | <0.001 | 0.69 | 0.73 |
| Age (decade)* | 0.84 | <0.001 | 0.84 | 0.85 |
| Race (White) | 0.93 | <0.001 | 0.91 | 0.96 |
| anxiety | 0.80 | <0.001 | 0.77 | 0.82 |
| depression | 0.64 | <0.001 | 0.61 | 0.67 |
| psychosis | 0.63 | <0.001 | 0.56 | 0.71 |
| suicide | 0.73 | <0.001 | 0.66 | 0.81 |
| violence | 0.40 | <0.001 | 0.37 | 0.43 |

## Discussison

In this EHR study, we detect marked reduction in notes (including those from telemedicine) documenting psychiatric symptoms, paralleling the emergence of COVID-19 diagnoses in the Boston area. During this period outpatient visits were cancelled, individuals likely became more reluctant to come to the ED, and clinicians seeking to reduce exposure may have conducted more focused interviews. Conversely, among ED evaluations, while most symptoms were not associated with testing, reference to violence was associated with greater likelihood of testing, perhaps reflecting a shift in clinical presentations.

### Limitations

Limitations of these results include the reliance on simple string-matching, and the unclear generalizability from these 5 hospitals to other regions.

### Conclusions

Nonetheless, our results indicate an acute reduction in assessment and documentation of psychiatric symptoms. In light of survey data^2^, one might expect to see an increase rather than decrease in psychiatric presentations in Boston-area hospitals. Efforts to provide more accessible psychiatric care during the acute phase of the COVID-19 pandemic may become particularly important: symptoms are likely to be increasing, while access is objectively decreasing. Strategies such as telemedicine are urgently needed to ensure that another consequence of the pandemic is not neglect of psychiatric illness.

## Data Availability

Participant-level data has not been approved for release by the IRB.

## Acknowledgements

Dr. Perlis had full access to all the data in the study and takes responsibility for the integrity of the data and the accuracy of the data analysis.

No funding was received for this study. Dr. Perlis has received consulting fees from Burrage Capital, Genomind, RID Ventures, and Takeda. He holds equity in Outermost Therapeutics and Psy Therapeutics. Mr. Castro reports no conflict of interest.

## References

1. Brooks SK, Webster RK, Smith LE, et al. The psychological impact of quarantine and how to reduce it: rapid review of the evidence. Lancet Lond Engl. 2020;395(10227):912–920. doi:10.1016/S0140-6736(20)30460-8

2. New Poll: COVID-19 Impacting Mental Well-Being: Americans Feeling Anxious, Especially for Loved Ones; Older Adults are Less Anxious. https://www.psychiatry.org/newsroom/news-releases/snew-poll-covid-19-simpacting-mental-well-being-americans-feeling-anxious-especially-for-loved-ones-older-adults-are-less-anxious. Accessed March 27, 2020.

3. Lowell A, Suarez-Jimenez B, Helpman L, et al. 9/11-related PTSD among highly exposed populations: a systematic review 15 years after the attack. Psychol Med. 2018;48(4):537–553. doi:10.1017/S0033291717002033

4. Shalev AY, Gevonden M, Ratanatharathorn A, et al. Estimating the risk of PTSD in recent trauma survivors: results of the International Consortium to Predict PTSD (ICPP). World Psychiatry Off J World Psychiatr Assoc WPA. 2019;18(1):77–87. doi:10.1002/wps.20608

5. Nalichowski R, Keogh D, Chueh HC, Murphy SN. Calculating the benefits of a Research Patient Data Repository. AM1A Annu Symp Proc. 2006;2006:1044.

6. McCoy TH, Yu S, Hart KL, et al. High Throughput Phenotyping for Dimensional Psychopathology in Electronic Health Records. Biol Psychiatry. 2018;83(12):997–1004. doi:10.1016/j.biopsych.2018.01.011

